# Clinician Experiences with Ambient AI Scribe Technology in Singapore: A Qualitative Study

**DOI:** 10.64898/2026.03.17.26348627

**Authors:** Ravi Shankar, Aaron Goh, Qian Xu

## Abstract

**Background:** The administrative burden of clinical documentation is a recognised contributor to clinician burnout and diminished care quality. Ambient artificial intelligence (AI) scribe technology, which uses large language models to passively record and summarise clinical encounters, has rapidly gained traction internationally. However, no published studies have examined clinician experiences with this technology in the Asia-Pacific region or within Singapore’s multilingual healthcare system.

**Objective:** This study explored clinician perspectives on ambient AI scribe technology at Alexandra Hospital, Singapore, focusing on perceived benefits, barriers, workflow integration, ethical considerations, and recommendations for sustained implementation.

**Methods:** A qualitative descriptive study was conducted using semi-structured interviews with 28 clinicians across multiple specialties at Alexandra Hospital, National University Health System (NUHS). Participants were purposively sampled for diversity in role, specialty, and usage level. Interviews were analysed using reflexive thematic analysis guided by the RE-AIM/PRISM framework. The COREQ checklist was followed.

**Results:** Five themes emerged: (1) reclaiming presence in the clinical encounter, (2) navigating accuracy and trust in AI-generated documentation, (3) workflow disruption and adaptation, (4) privacy, consent, and ethical tensions within Singapore’s regulatory landscape, and (5) envisioning sustainable integration. Clinicians reported improved patient engagement and reduced cognitive burden. Persistent barriers included accuracy concerns, AI hallucinations, limited multilingual functionality, loss of documentation style, and uncertainties around compliance with the Personal Data Protection Act (PDPA).

**Conclusions:** Ambient AI scribe technology holds promise for alleviating documentation burden in Singapore’s public healthcare system. Realising this potential requires attention to safety validation, multilingual capability, clinician training, and patient-centred consent aligned with local regulatory frameworks.

## 1. Introduction

Clinical documentation has become one of the most time-consuming aspects of modern healthcare delivery. Physicians spend approximately two hours on electronic health record (EHR) tasks for every hour of direct patient care, with after-hours documentation contributing substantially to professional dissatisfaction and burnout [1,2]. This documentation burden has been identified as a primary driver of the clinician burnout epidemic, affecting patient safety, care quality, and workforce retention [3,4].

Ambient artificial intelligence (AI) scribe technology has emerged as a promising innovation to address this challenge. These tools passively capture clinician-patient conversations, leverage large language models (LLMs) to transcribe and summarise encounters, and generate structured draft clinical notes for clinician review [5,6]. Unlike traditional dictation software, ambient AI scribes operate unobtrusively, enabling clinicians to maintain natural engagement with patients while documentation occurs in the background [7].

Adoption has accelerated rapidly. By 2025, approximately two-thirds of physicians in the United States reported using AI tools in clinical practice [8]. Major health systems, including Kaiser Permanente and the US Veterans Affairs system, have deployed ambient AI scribes at enterprise scale [9,10]. Early evidence suggests time savings of 20% to 30% in documentation, with a landmark randomised clinical trial of 238 physicians demonstrating reduced writing time, lower burnout, and decreased task load [11,12,13].

However, concerns persist regarding the safety and accuracy of AI-generated clinical notes. Modern LLM-based ambient scribes report error rates of approximately 1% to 3%, including hallucinations, critical omissions, and contextual misinterpretations [14]. Recent editorials have argued that adoption is outpacing validation, with the field’s focus on efficiency metrics potentially overshadowing questions of clinical safety [15,16].

Qualitative understanding of clinician experiences remains limited. Existing studies have focused on physician perspectives in North American and Australian settings, with little representation of non-physician clinicians or healthcare systems in the Asia-Pacific region [17,18,19]. Singapore’s multilingual clinical environment, distinct regulatory frameworks including the Personal Data Protection Act (PDPA), and integrated public healthcare delivery model present implementation considerations that differ substantively from those reported in the existing literature. No published study has examined ambient AI scribe implementation in this context.

This study addresses this gap through a qualitative exploration of clinician experiences with ambient AI scribe technology at Alexandra Hospital, a tertiary hospital within the National University Health System (NUHS) in Singapore.

## 2. Literature Review

### 2.1 Documentation Burden and Clinician Burnout

Electronic health records, while delivering benefits in legibility, data accessibility, and interoperability, have imposed substantial documentation demands. Documentation-related tasks consume a disproportionate share of working hours, with additional after-hours charting, termed “pajama time,” compounding the problem [1,2,20]. The cumulative effect includes burnout, reduced professional fulfilment, impaired patient-clinician communication, and downstream effects on care quality [3,4].

### 2.2 Evolution of Ambient AI Scribes

Clinical documentation technology has progressed from dictation software to human scribes to AI-powered ambient tools. Modern ambient AI scribes use LLM-based natural language processing to generate structured notes from passively captured conversational audio [5,6]. Commercial platforms including Microsoft DAX Copilot, Abridge, Nabla, and Suki now compete in a marketplace that has attracted over US$600 million in venture capital [21,22]. The technology has been described as potentially the fastest digital technology adoption in healthcare history [15].

### 2.3 Evidence on Effectiveness and Safety

A rapid review of real-world evidence identified reductions in documentation time and modest decreases in after-hours work, alongside improvements in clinician satisfaction [23]. A quality improvement study of 46 clinicians found that ambient scribing reduced time in notes by 20.4% per appointment and after-hours work by 30.0% [11]. A randomised trial of 238 physicians across 14 specialties confirmed reductions in writing time and burnout with two different ambient AI platforms [13].

Safety evidence is more equivocal. Validated quality assessments have found AI-generated notes to be more thorough but less succinct and more prone to hallucination than physician-authored notes [24]. Error rates of 1% to 3% are reported, though even low rates carry serious implications in clinical documentation [14]. Concerns about long-term “cognitive debt” from overreliance on AI documentation have also been raised [16,25].

### 2.4 Qualitative Evidence and Gaps

Shah et al. (2025) conducted 22 semi-structured interviews with physicians at Stanford, guided by the RE-AIM/PRISM framework, finding positive impacts on workload and patient engagement alongside barriers including limited multilingual functionality and accuracy concerns [17]. Van Tiem et al. (2026) interviewed 24 clinicians at the University of Iowa, identifying tensions around note formatting and loss of personal documentation voice [18]. Focus groups with 33 Australian medical practitioners found insufficient AI knowledge, medico-legal risks, and privacy concerns as barriers [19]. A mixed-methods pediatric study found only 32% encounter utilisation and heterogeneous experiences [26].

No published qualitative study has examined clinician experiences with ambient AI scribes in the Asia-Pacific context. This study provides the first such exploration, situated within Singapore’s unique multilingual, regulatory, and healthcare delivery environment.

## 3. Methods

### 3.1 Study Design

A qualitative descriptive study was conducted [27], guided by the RE-AIM/PRISM framework [28]. The Consolidated Criteria for Reporting Qualitative Research (COREQ) 32-item checklist was followed [29].

### 3.2 Setting

The study was conducted at Alexandra Hospital (AH), a tertiary hospital within the National University Health System (NUHS) in Singapore. Alexandra Hospital provides a comprehensive range of medical services to a diverse patient population in the southwestern region of Singapore. The hospital had implemented an enterprise-wide ambient AI scribe platform across outpatient clinics, selected inpatient wards, and the emergency department, available to physicians, advanced practice nurses, and allied health professionals. At the time of data collection, the platform had been available institution-wide for approximately six months, preceded by a three-month pilot.

Singapore’s multilingual patient population, which includes English, Mandarin, Malay, Tamil, and various dialect speakers, creates distinct challenges for AI-based clinical documentation. The regulatory landscape, governed by the Personal Data Protection Act (PDPA) 2012 and the anticipated Health Information Bill (HIB), imposes specific requirements on personal health information handling.

### 3.3 Participants

Purposive sampling was employed to recruit participants with diversity in clinical role, specialty, practice setting, and usage level [30]. Twenty-eight clinicians participated: 18 physicians (64.3%), 6 advanced practice providers (21.4%), and 4 allied health professionals (14.3%), representing 11 specialties. Ten participants (35.7%) were high users (more than 70% encounter activation), nine (32.1%) moderate users (30% to 70%), five (17.9%) low users (less than 30%), and four (14.3%) had discontinued use. Recruitment continued until thematic saturation was achieved [31].

### 3.4 Data Collection

Semi-structured interviews were conducted between January and April 2026 via a secure video conferencing platform. An interview guide was developed from the RE-AIM/PRISM framework and adapted for the Singapore context through pilot testing with three clinicians (excluded from analysis). Domains included: motivations and expectations, practical experiences, documentation quality, patient-clinician interaction, accuracy and safety, privacy and regulatory compliance, multilingual encounters, and recommendations. Interviews lasted 30 to 55 minutes (mean: 41 minutes), were audio-recorded with consent, and transcribed verbatim. Field notes were maintained by the interviewing researcher (RS).

### 3.5 Data Analysis

Reflexive thematic analysis was conducted following Braun and Clarke’s six-phase approach [32]. Three researchers (RS, AG, XQ) independently coded a subset of transcripts to develop the initial codebook, with consensus achieved through iterative discussion. Coding was performed using Dedoose (version 9.2; SocioCultural Research Consultants, Los Angeles, CA). An audit trail was maintained throughout.

### 3.6 Ethics

Ethical approval was obtained from the National University Health System Domain Specific Review Board (NHG DSRB). The study was conducted in accordance with the Declaration of Helsinki and the Human Biomedical Research Act (HBRA). Written informed consent was obtained from all participants. Data management complied with NUHS governance policies and the PDPA.

## 4. Results

Five themes emerged from the analysis, presented below with illustrative quotations.

### 4.1 Reclaiming Presence in the Clinical Encounter

The most consistently reported benefit was the restoration of meaningful presence during clinical encounters. Clinicians described a shift in their ability to engage with patients when freed from simultaneous documentation demands.

A general medicine physician (P3) reflected: “For the first time in years, I can actually look my patients in the eye during the entire visit. I am not typing, I am not glancing at the screen. I am listening.” A nurse practitioner (P19) observed: “Our patients have commented that consultations feel different now. They feel heard. That is incredibly meaningful, especially for our elderly patients.” This observation holds particular resonance in Singapore’s ageing society, where effective communication with older adults requires sustained attention and culturally sensitive engagement.

This benefit was less pronounced in procedure-heavy specialties. A general surgeon (P8) noted: “The ambient scribe does not really help with operative notes. We have highly specific templates that need exact measurements and findings.” A rehabilitation medicine physician (P24) similarly found that follow-up visits involving structured functional assessments were less amenable to ambient documentation.

### 4.2 Navigating Accuracy and Trust

AI hallucination was the most concerning accuracy issue. An internist (P5) described discovering a fabricated medication entry: “It listed atorvastatin as a current medication. The patient has never been on atorvastatin.” A geriatrician (P15) recounted a case where symptoms were misattributed to the patient rather than a family member discussed during the encounter.

A psychiatrist (P14) noted: “In mental health, nuance is everything. The scribe captured the factual content but missed the emotional context. The distinction between a patient saying they have thought about death versus actively planning self-harm is enormous, and the AI does not always capture that distinction.”

An emergency physician (P22) remarked: “The notes read like they were written by a trainee. They are technically correct but lack the clinical judgement and prioritisation that an experienced physician brings.” This was troubling for clinicians who viewed notes as clinical communication with future care providers across Singapore’s integrated care model.

A family medicine physician (P2) observed: “I spend less time writing, but I spend more time reading and correcting. Some days I am not sure the net time savings are as large as they initially seemed.”

### 4.3 Workflow Disruption and Adaptation

Ambient AI scribes were most beneficial in outpatient settings. In inpatient wards, the technology fit less naturally. A medical officer (P17) explained: “On the inpatient side, the pre-templated formats we already use are very streamlined, and adding an AI-generated layer creates more work rather than less.”

This theme was of particular relevance to the Singapore context. An internist (P6) described: “Many of my older patients speak primarily in Mandarin or Hokkien. When the conversation switches from English, the accuracy drops significantly, and mixed-language passages are sometimes garbled entirely.” A family medicine physician (P2) added: “For my Malay-speaking patients, I sometimes have to rewrite the note almost from scratch. The tool was clearly not designed for our multilingual reality.”

Clinicians described modifying their conversational behaviour to optimise scribe performance, including verbalising clinical reasoning and adopting more structured dialogue patterns. Some viewed this positively; others perceived it as constraining natural interaction. A psychiatrist (P14) noted: “I find myself narrating my thought process out loud, which changes the dynamic of the therapeutic encounter.”

### 4.4 Privacy, Consent, and Ethical Tensions

Several clinicians expressed uncertainty about whether patients truly understood the implications of ambient AI documentation. A geriatrician (P15) raised concerns about consent capacity: “Some of my patients with early dementia may not fully understand what they are consenting to.”

A senior consultant (P10) remarked: “The PDPA requires that we collect personal data only for purposes that a reasonable person would consider appropriate. I am not entirely sure all our patients would consider AI recording of their consultation to be within their expectations.” Questions about data residency, cloud processing, and third-party vendor involvement were raised by multiple participants.

A psychiatrist (P14) noted: “Patients sometimes share information they do not want in the medical record. With the AI scribe running, everything is captured, and I have to actively edit things out.” This concern was compounded by cultural considerations around disclosure of sensitive personal and family matters.

### 4.5 Envisioning Sustainable Integration

Clinicians with the highest satisfaction consistently cited robust training, peer support, and ongoing technical assistance. A general practitioner (P1) emphasised: “The training was essential. Having a colleague show me how to customise the templates made all the difference.” Those with minimal onboarding were more likely to discontinue.

The ability to personalise note templates and adjust detail levels was identified as critical. An orthopaedic surgeon (P10) suggested: “If I could instruct the system to generate a brief assessment for follow-ups but a detailed history for new patients, that would make it much more useful.”

A nurse practitioner (P20) noted: “Hearing from colleagues about how they use it and the tricks they have discovered was more helpful than any formal training session.” Clinical champions and communities of practice were viewed as essential for sustained adoption.

An internal medicine physician (P6) recommended: “There should be a structured process for reporting errors and hallucinations, with regular audits of note accuracy.” Participants suggested leveraging Alexandra Hospital’s existing clinical governance infrastructure for this purpose.

**Figure 1.**
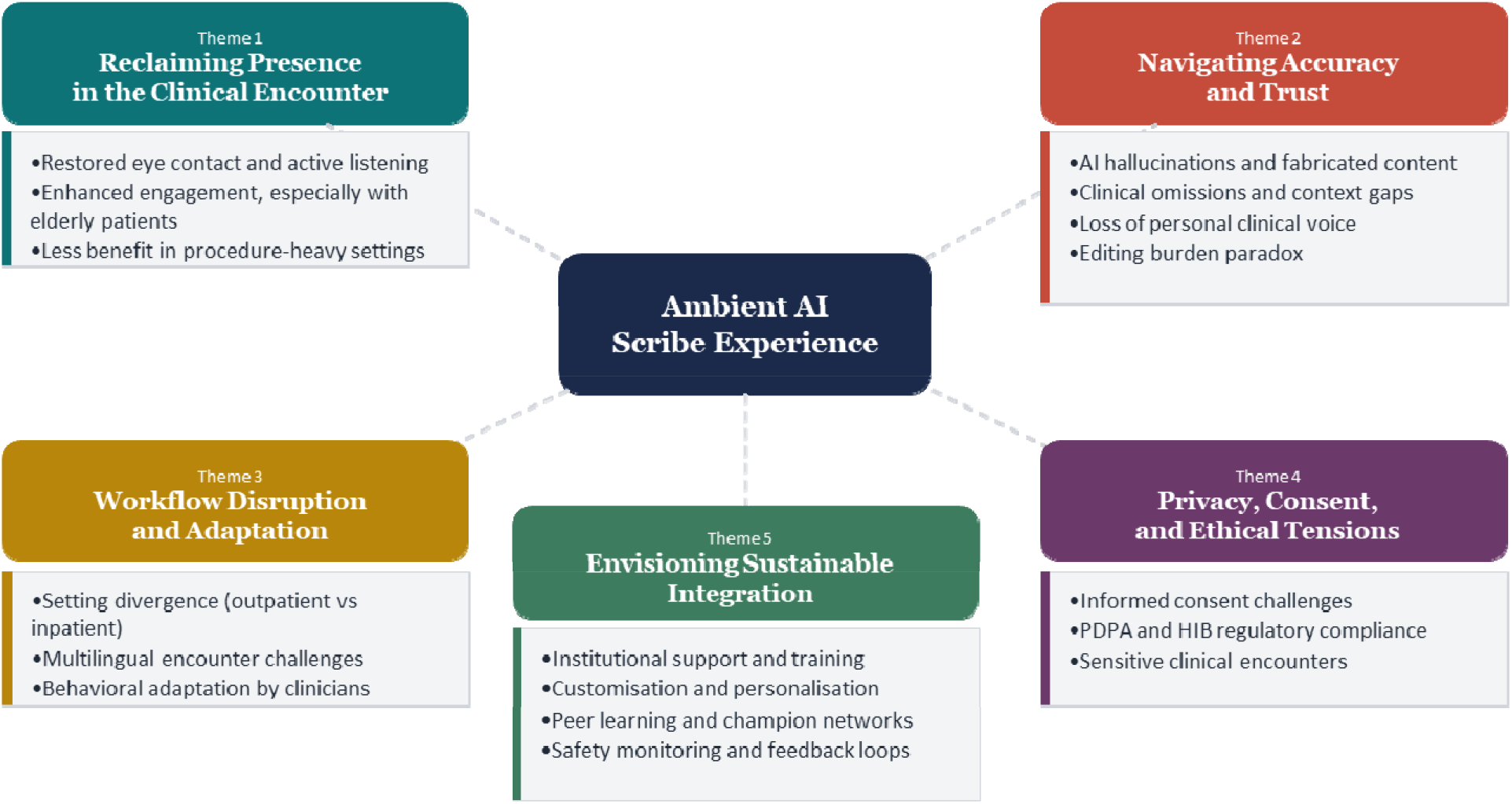
Thematic Map of Clinician Experiences with AI-Powered Ambient Scribe Technology

## 5. Discussion

### 5.1 Principal Findings

This study provides the first qualitative exploration of ambient AI scribe implementation in the Asia-Pacific healthcare context. The most prominent finding is the transformative impact on the quality of the clinician-patient interaction, consistent with prior research [17,18,19]. Clinicians at Alexandra Hospital described a fundamental shift in their ability to be present with patients. This finding is especially significant in Singapore, where communicating with a linguistically and culturally diverse, ageing patient population makes encounter quality particularly important.

Concerns regarding accuracy, hallucination, and documentation quality are consistent with the international literature [14,24]. This study adds depth by demonstrating that these issues carry relational and professional dimensions: the loss of clinical voice, the editing burden paradox, and the challenge of capturing nuance all reflect the complex relationship between clinicians and their documentation practices.

The identification of multilingual encounter challenges as a major barrier is a novel contribution. While prior studies have noted limited functionality with non-English-speaking patients [17], this study documents the specific challenges of code-switching, dialect diversity, and mixed-language encounters that characterise healthcare delivery in Singapore. This finding has equity implications, as patients communicating predominantly in non-English languages may receive lower quality AI-generated documentation.

### 5.2 Implementation Perspectives

Viewed through the RE-AIM/PRISM framework, the technology’s reach was moderated by language capabilities and specialty suitability. Effectiveness was perceived positively for time savings and patient engagement but tempered by accuracy concerns. Adoption was facilitated by ease of use and peer support but hindered by multilingual limitations and privacy concerns. Implementation success depended on training quality and EHR integration. Maintenance highlighted the need for sustained safety monitoring and institutional investment.

### 5.3 Implications for Practice

Several practical implications emerge. First, deployment should be staged and specialty-sensitive, prioritising outpatient, conversation-driven specialties. Second, training programmes incorporating peer learning and clinical champions are essential. Third, consent processes must be transparent, with particular attention to capacity in cognitively impaired populations and alignment with the PDPA. Fourth, systematic safety monitoring should be integrated into existing clinical governance frameworks. Fifth, investment in multilingual AI capabilities accommodating Singapore’s linguistic diversity is urgently needed.

### 5.4 Implications for Research

Longitudinal studies are needed to assess sustained benefits. Patient perspectives on AI-documented encounters remain underexplored, particularly in the Asian context. Research on clinical de-skilling, cost-effectiveness, and multilingual ambient AI performance in Asian languages are priorities. Comparative studies of implementation strategies across different healthcare systems would also be valuable.

### 5.5 Strengths and Limitations

Strengths include this study’s position as the first qualitative exploration of ambient AI scribes in Singapore, the inclusion of diverse clinical roles, multiple specialties, use of an established implementation science framework, and adherence to COREQ. The identification of multilingual challenges is a novel contribution.

Limitations include the single-site design at Alexandra Hospital, which may limit transferability. The self-selected sample may introduce bias. The cross-sectional design does not address how experiences evolve. Patient perspectives were not captured and represent an important area for future research.

## 6. Conclusions

Ambient AI scribe technology holds substantial promise for addressing the documentation burden in Singapore’s public healthcare system. This study demonstrates that the technology’s most valued contribution is the restoration of meaningful human presence in the clinical encounter. However, sustainable integration requires systematic attention to accuracy validation, multilingual functionality, regulatory compliance, and clinician training. The heterogeneity of clinician experiences underscores the need for differentiated, specialty-sensitive implementation strategies. These findings from Alexandra Hospital provide a foundation for evidence-informed deployment across Singapore’s healthcare landscape and offer a model for evaluating ambient AI scribe adoption in healthcare systems across the Asia-Pacific region.

## Data Availability

All data produced in the present work are contained in the manuscript

